# Advancing convergent mixed methods design using the Diamond Approach for clinical multiple case studies: an example using data on time to treatment in aneurysmal subarachnoid haemorrhage

**DOI:** 10.1101/2021.09.08.21263308

**Authors:** Thuy Phuong Nguyen, Christine Stirling, Gemma Kitsos, Kim Jose, Linda Nichols, Ronil V. Chandra, Sabah Rehman, Karen Smith, Ian Mosley, Leon Lai, Hamed Asadi, Arvind Dubey, Jens Froelich, Amanda Thrift, Seana Gall

## Abstract

Using a new approach to the analysis and synthesis of a clinically focused mixed-methods multiple case study of aneurysmal subarachnoid haemorrhage (aSAH), we explored barriers and facilitators to timely treatment. In this paper we provide guidance for the multiple steps of analysis and synthesis of qualitative and quantitative data from across and within 27 case studies. Results showed that median (IQR) time to treatment was 15.1 (9.0, 24.1) hours. Only 37% of cases had treatment within 12-hours of onset. Early recognition of aSAH, good coordination, and availability of resources for treatment were main facilitators for treatment within 12 hours from onset. Lack of recognition of aSAH at onset and lack of resources for immediate in-hospital treatment were major barriers.

## Introduction

Aneurysmal subarachnoid haemorrhage (aSAH) is a medical emergency that requires early treatment through surgical clipping or endovascular coiling to achieve better outcomes for patients (Connolly et al., 2012; Phillips et al., 2011). While there is no consensus about optimal treatment times, receiving treatment within 12 hours (Buscot et al., 2021) or 24 hours (Phillips et al., 2011) is associated with fewer complications, greater discharge to home, reduced disability and greater survival than treatment after those periods. Buscot et al. (2021) demonstrate that receiving aSAH treatment within 12.5 hours led to greater odds of being discharged home than being discharged to rehabilitation regardless of severity, gender, treatment type and transfer. Phillips et al. (2011) show that surgery at ≤ 24 hours compared to >24 hours was associated with a 44% (95% CI 69%, 99%) relative risk reduction of death or disability 6 months later. Ensuring that people with aSAH are able to access treatment at a specialised facility as soon as possible after symptom onset is important to achieve the best possible outcomes.

Delayed treatment for aSAH appears to be common. In one study from a single hospital in Melbourne, only half the people with aSAH were treated within 24 hours of onset (Phillips et al., 2011) while in a country-wide study in the UK more than 60% were treated >2 days after onset of symptoms (Langham et al., 2009) and in the US 10–25% of people are treated at >48 hours after admission (Sarmiento et al., 2015; Siddiq et al., 2012). Our understanding of the causes of these delays remains limited, in turn affecting our ability to develop interventions targeted at reducing delays and improving outcomes.

In a systematic review of predictors of time to treatment of aSAH, most predictors (i.e. being female, older age, living in urban areas, having inter-facility transfer) had a bidirectional influence on time to treatment (Nguyen, Rehman, et al., 2021). Other potential predictors of delays in aSAH treatment include pre-hospital delays (Germans et al., 2014), admission on the weekend (Sarmiento et al., 2015; Siddiq et al., 2012), having pre-treatment complications (Sonig et al., 2018) or having surgical clipping (Sarmiento et al., 2015; Sonig et al., 2018). None of these indicators clearly predicted longer or shorter time to treatment after aSAH. Moreover, a critical problem with prior studies is the use of secondary quantitative databases that include limited relevant contextual data, thus hindering the potential to fully explore reasons for delay (Nguyen, Rehman, et al., 2021). Nguyen et al (2021) also found that studies were focussed on single time periods of the patients’ pathway to treatment, or on single centres, and these are not representative of the patients’ journey across the continuum of healthcare service provision.

To date, quantitative studies have produced evidence on “what works’, but these studies often fail to answer the question “why it works/does not work”. To answer such questions, complexities and intricacies of programs, people, and places must be taken into consideration (Padgett, 2012) as well as the socio-economic, cultural and environmental contexts (Dahlgren & Whitehead, 2006), which could be studied effectively using qualitative research. Qualitative research seeks in-depth understanding of social phenomena and patients’ perspectives on health and illnesses, and health services (Pope & Mays, 2020). Further it highlights the role of human and system interactions within complex health systems, thereby generating new insights for health interventions and policy. The addition of qualitative data to aSAH studies can improve our ability to understand and address the delays in treating aSAH. An in-depth exploration of the context of aSAH events and the decisions made in the process of seeking and providing care using a case study approach may improve our understanding of the bi-directional nature of predictors of time to treatment.

Combining quantitative and qualitative methods enhances understanding of the phenomenon of interest by facilitating completeness in data and triangulation of results (Greene, 2007). Social science researchers first used triangulation technique to validate their research results (Mertens & Hesse-Biber, 2012). Triangulation employs multiple methods, investigators, theories or data sources to gain multiple perspectives and reduce the chance of reaching false conclusion (Carter et al., 2014). Triangulation is also used to provide complementary information on the same phenomenon (Hammersley, 2008). This does not contradict with the primary validation purpose of triangulation as complementary information could correct initial interpretation of data and shed new understanding of the phenomenon. Although mixed methods approaches have been used in medical research for decades (Regnault et al., 2018), challenges in analysing and synthesising data in these studies remain. Casey et al. (2016), who combined patients’ clinical data with qualitative data to assess the outcomes from a diabetes related intervention study, identified that the lack of published guidance on how to merge and interrogate data in a concurrent triangulation study was a limitation (Casey et al., 2014).

We, therefore, aimed to use a mixed-methods multiple case study approach to explore thoroughly the facilitators and barriers to timely treatment of aSAH. Our article also provides guidance for a new approach to the analysis and synthesis of data from a clinically-focused multiple case study through the process of answering our empirical research questions. Our research questions were: (1) What are the significant factors influencing time to treatment of aSAH patients? (2) How much do the factors influence time to treatment of aSAH patients?

## Methods

### Design

We used a convergent mixed-method multiple case study approach with a novel crossover analysis to answer our research questions. We believe this answers Hitchcock & Onwuegbuzie’s (2019) call to innovate new crossover analysis for mixed methods clinical research. Our pragmatic *Diamond Approach to Mixed Methods Multiple Case Studies* (herein “The Diamond Approach”) provides comprehensive guidance to a crossover analysis and synthesis of data from a clinically focused mixed-methods multiple case study. Applying Hitchcock & Onwuegbuzie’s framework (2019), our study can be described as a crossover mixed analysis approach where pragmatism and cases studies formed the basis for mixing the qualitative and quantitative methods (Hitchcock & Onwuegbuzie, 2019). The Diamond Approach’ techniques described here integrated qualitative and quantitative data from across and within case studies in multiple steps of varying complexity (Figure 1).

**Figure 1.**
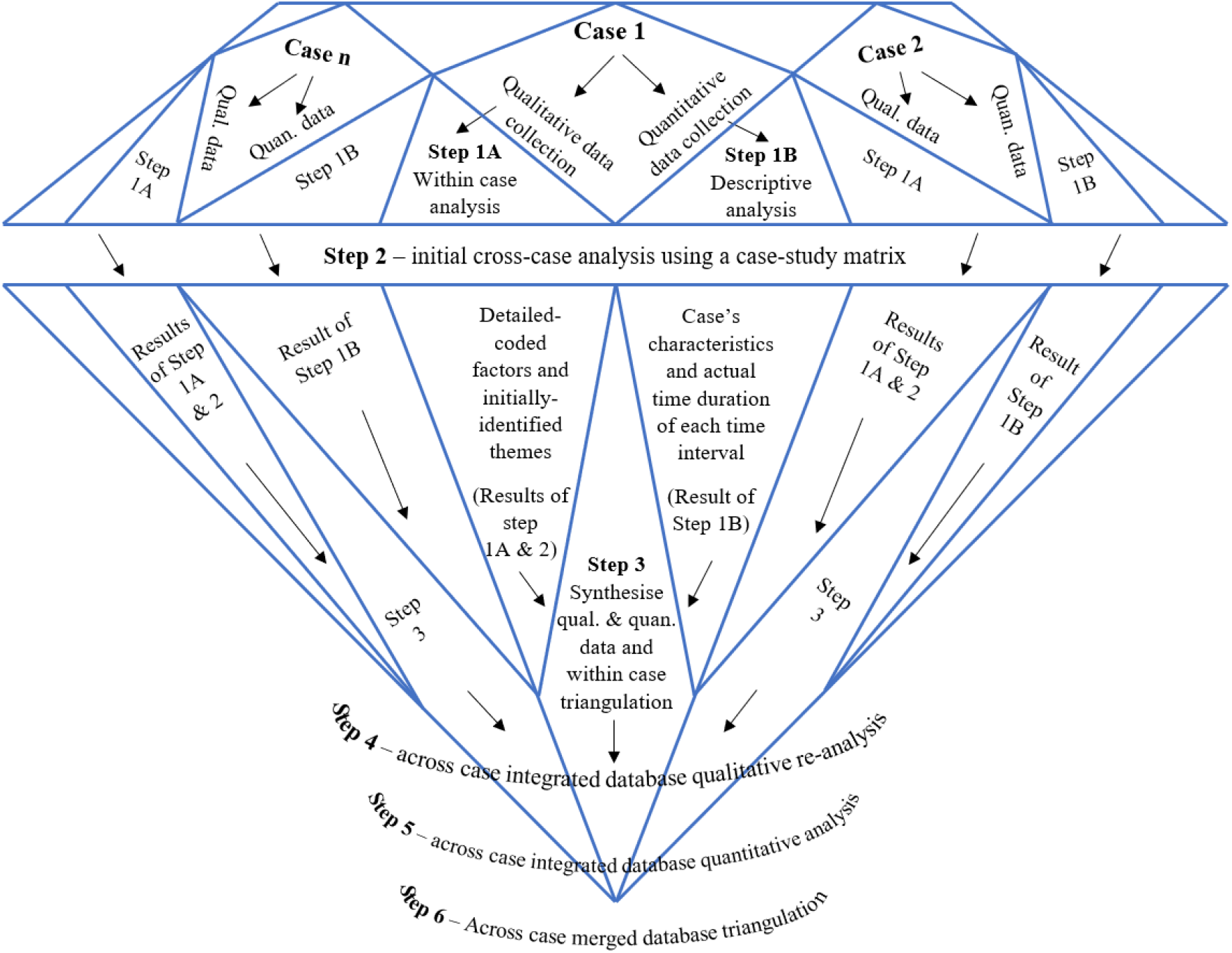
The *Diamond Approach to Mixed Methods Multiple Case Studies*

### The Case

We defined a case as one episode of aSAH being hospitalised at one of two tertiary hospitals in Australia. The participant with aSAH was at the centre of each case study, with any key individuals (such as a spouse or friend) and health professionals involved in the event also included. We collected relevant data from hospital administrative, patient medical and ambulance records. Cases were bounded by the start of the event (onset of aSAH) until the patient was treated with either endovascular coiling or surgical clipping. Cases were identified prior to data collection following a deductive approach (Creswell & Creswell, 2018).

We developed a two-dimensional *a priori* data framework consisting of major time intervals along the patient’s pathway for thematic coding. For each factor identified, coding indicated whether the impact on time to treatment was positive or negative to timely treatment. We defined five main time points during the patient’s pathway as i) onset of event, ii) arrival to first hospital, iii) receiving first diagnostic computer tomography (CT), iv) arrival to tertiary hospital (for patients who were transferred) and v) receiving definitive treatment. The time from symptom onset of aSAH until the patient arrived at the first hospital was labelled as the “pre-hospital” interval. From first hospital arrival until the patient received first CT was labelled as the “diagnostic” interval. Interfacility transfer time between first hospital and the tertiary hospital was labelled as the “transfer” interval (patient might be transferred through several hospitals until they reached the treating hospital). The “in-hospital” interval was determined as the duration between first CT to definitive treatment. For patients undertaking an interfacility-transfer, the transfer time is contained within the time block. A factor that prolonged any time interval was defined as a “barrier”. In contrast, a factor that facilitated shorter time to treatment was identified as a “facilitator”.

### Setting

Our study was conducted within the referral networks of two tertiary hospitals in South East Australia. Both institutions are tertiary hospitals with capacities to treat aSAH in their respective regions including neurosurgery, interventional neuroradiology and intensive care units. Hospital A. covers an area with a population of about 1.5 million and Hospital B. provide services to a population of 500,000. With an incidence of aSAH of 8.5/100,000 people per year (Etminan et al., 2019), we anticipated approximately 170 cases per year.

### Participants

We recruited patients with aSAH who survived >1 day after an aSAH. Immediately catastrophic cases were unlikely to inform our aims and were therefore excluded. Hospital admission and discharge lists from March 2019 to March 2020 were prospectively screened to identify cases of aSAH (consent for the case finding was waived). Consent to participate in the study was provided by either the patient if they were able to give consent or by patient’s next-of-kin (NOK). In total we recruited 27 patients. Using cases identified only inside the healthcare system did not affect the representativeness of data since people with aSAH are almost all hospitalised (Nichols et al., 2020).

Once the patient consented, we used an active snowball recruitment technique (Patton, 2002) to recruit the person (called ‘key individual’) who was with the patient at the time of the event or first on the scene. In our study, all key individuals present at the event were NOK. We reviewed the patient’s notes and/or the ambulance database to identify the health professionals involved in the management of the patient. We recruited paramedics (11), emergency physicians (5), interventional neuroradiological consultants or fellows (15), neurosurgical consultants or registrars (27) and theatre nurses (1) into the study. Written informed consent was obtained from all health professionals. We used an interview tree (Supplement 1) to monitor the process of recruiting and interviewing participants. Participation rate of health professionals was approximately 40%.

### Qualitative instruments and data collection

For qualitative data, we used an in-depth interview method. Following identification and consent by people with aSAH and relevant stakeholders involved in the case, a semi-structured face-to-face interview was conducted with 90 interviewees. Questions for patients and/or their NOK focused on the events between the onset of symptoms and arrival at hospital, including factors that influenced their decision-making. For paramedics, questions focused on perceived barriers and facilitators to the person reaching the treating hospital in a timely manner, including signs or symptoms present, aspects of care provided at the scene or hospital factors, such as ambulance ramping or bypass. Questions for health professionals focused on the events surrounding diagnosis and treatment.

We used NVivo 12 Qualitative Data Analysis Software by QSR International to manage and analyse qualitative data (QSR International, 1999). All interviews related to a patient with aSAH became part of the data for the same case (herein “the Case”). The number of interviews differed from case to case, ranging from 2 – 5 interviews, depending on factors such as consent to participate, and the number of individuals involved in the case.

### Quantitative instruments and data collection

We extracted data on individual factors, pre-hospital factors, and hospital factors from the digital medical records for all eligible people with aSAH who consented to participate in the study. Each patient with aSAH was also linked to the dispatch and clinical databases of ambulance services (if applicable) to collect ambulance related data. Data were extracted using a pre-designated form on REDCap electronic data capture tools hosted at University of Tasmania (Harris et al., 2019), already in use in a larger study (Buscot et al., 2021), by research assistants trained in the procedure. Data collected included date and time of all available events from the onset until patient received definitive treatment (onset, ambulance called, ambulance arrival, hospital arrival, triage time, CT/MRI/lumbar puncture time, transfer time, procedure time), and demographic information (age, sex, marital status, ethnicity, insurance status and address details for linkage to spatial social disadvantage and remoteness data), medical history (vascular risk factors, lifestyle factors, and medication history), pre-hospital care (time of call, dispatch code, case nature, assessments including vital signs, scene arrival and departure times, distances travelled, transport method, destination, reasons for choosing destination, and pre-existing conditions), clinical details (presenting symptoms and signs, preliminary diagnosis, the location and size of the aneurysm, and World Federation of Neurosurgeons Scale (WFNS) with value from 1 to 5 with higher grade predicts more severe aSAH symptoms), and treatment method.

### Mixed methods data analysis, integration and synthesis

#### Concurrent Step 1A – within case qualitative analysis

We used a “codebook” thematic analysis approach (Braun & Clarke, 2020), in which a structured coding framework was used to develop and document the analysis. Each interview transcript was coded using the *a priori* framework to identify when that factor occurred along a patient’s pathway to treatment and whether a factor was a barrier or facilitator to more timely treatment.

Next, we undertook detailed coding of the factors (either barriers or facilitators) identified in the initial coding process based on a conceptual framework on health care access (Levesque et al., 2013). We also allowed new factors to emerge during this coding phase. All factors were categorised within the relevant time interval.

#### Concurrent Step 1B – within case quantitative analysis

Quantitative data were analysed separately using R 4.0.3 (R Core Team, 2020). We calculated time duration for each interval in one patient’s pathway to treatment, in accordance with the time interval in the qualitative analysis framework (pre-hospital, diagnosis, transfer and in-hospital intervals). Time duration for each interval was determined from dates and times of five major time points extracted from ambulance, emergency department, radiology and/or surgical records. In addition, we calculated time from onset to ambulance call, time from ambulance call to ambulance arrival. We performed descriptive analysis of patients’ demographic and clinical characteristics.

#### Step 2 – cross case qualitative analysis

Following the within-case analysis, we conducted an initial cross-case analysis using a case-study matrix. We searched among detailed-coded factors within each time interval across cases to identify similarities or patterns in multiple cases (Miles et al., 2014; Yin, 2006). We identified five broader themes from cases stretching across all time intervals (Nguyen, Stirling, et al., 2021). We used word cloud technique (McNaught & Lam, 2010) to demonstrate the importance of each theme across time intervals. A word cloud is a visualisation of texts in which the more frequently appeared keywords are presented more prominently in relation to others. We tabulated the frequency of theme keywords that appeared across cases as either facilitator or barrier in each time interval, i.e., into the *a priori* framework. The frequency of each theme in each cell of the *a priori* framework was translated into the font size of the keywords, with bigger font indicated greater frequency or more dominance. The word cloud for this step of analysis has been presented elsewhere (Nguyen, Stirling, et al., 2021).

#### Step 3 – synthesise qualitative & quantitative data and within case triangulation

We merged quantitative data (characteristic of aSAH cases and major time to treatment duration) into the qualitative database in NVivo. Quantitative data were assigned as “Attributes” in NVivo.

This integrated database allowed the research team to triangulate within-case quantitative and qualitative data. The team examined both the detail-coded factors and the themes initially identified against patients’ characteristics and actual time duration of each intervals. We examined all thematic factors identified in each time interval and compared it to the quantified time duration for that interval. We calculated median and interquartile range for each time interval and stratified by patient’s characteristics. Quantified time durations for each case were compared with the median duration of that time interval and/or with service benchmarks (if available) to determine if the patient experienced delays. Factors that did not appear to impact on the overall time to treatment in each period were removed. For example, an interviewee might have indicated that ambulance took a long time to arrive. However, by triangulating the actual time from the ambulance call to ambulance arrival, which was within the normal service benchmarks, we could attest ambulance service did not actually increase delays. The triangulation process also enabled the investigators to thoroughly validate qualitative coding results. That process could be referred to as the inside-outside legitimation method, in which *emic viewpoint* or interviewee’ perspective (insider) is challenged and integrated with *etic viewpoint* or objective quantitative data (outsider) to accurately reflect reality (Onwuegbuzie & Johnson, 2006). We revisited transcripts of cases with extremely long or short times for any of the stages of the treatment journey to find any factors that may have been missed.

#### Step 4 – across case integrated database qualitative re-analysis

After refining and confirming the major influencing factors on time to treatment of aSAH, we revised our key themes across all cases using the word cloud technique described in Step 2 (Supplement 2). This across-case triangulated revision identified the most important barriers and facilitators across each major time interval. This process reduced the number of key factors from five to three thereby increasing clarity about the most important factors.

#### Step 5 – across case integrated database quantitative analysis

Next, we ran matrix coding queries within the single integrated database between significant themes (barriers/facilitators), major time intervals (pre-hospital, diagnosis, transfer, and in-hospital), and case attributes (sex, age groups, early/delayed treatment, transferred/non-transferred groups, etc.) to identify patterns. As suggested from most recent literature, having treatment within 12 hours from onset of aSAH has shown to improve outcomes (Buscot et al., 2021; Kaneko et al., 2019). Hence, we used a 12-hour cut-off to classify patients receiving early/delayed definitive treatment for aSAH.

#### Step 6 – across case merged database triangulation

In our final step we triangulated the synthesised data to allowed identification of the density of each theme in each time interval stratified by early (<12 hours) or delayed (≥ 12 hours) treatment of aSAH. Again, we used word clouds to demonstrate the importance of facilitators and barriers to timely treatment of aSAH.

### Ethical considerations

Ethics approval was granted by Monash University (HREC/44916/MonH-2018-155101(v1)), Monash Health (RES-18-0000-648A) and University of Tasmania (H0017834). All participants provided informed consent before any data collection activities. Participants could opt out of the study at any time. Investigators were trained to work with vulnerable groups, ensuring our study met strictest ethical standards. We screened all potential cases with an aphasia and cognitive screening test to ensure they were able to provide informed consent. If a potential case did not have the ability to give informed consent, their next of kin was asked to consent (19 cases).

## Result

Our results include a summary of study participants and the key barriers and facilitators resulted from the triangulated, synthesised and integrated data in each stage of the patient journey. Quotes were attributed with their assigned case number, relationship to the person with aSAH. Any names of towns and hospitals had been removed from quotes to protect anonymity.

### Characteristic of aSAH cases

A total of 27 cases were included in our study, with 74% female. Age ranged from 24 to 87 years with almost half of all cases younger than 60 years of age. Most patients presented with headache (88.9%) while half of them experienced change in level of consciousness (48.1%). About 30% of patients presented with more severe conditions (WFNS grade 4 or 5). About 60% of cases received coiling as the definitive treatment. The median time from onset to either coiling or clipping was 15.1 hours (IQR 9.0 – 24.1 hours). More than one-third of patients had treatment within 12 hours of onset (Table 1).

**Table 1.** Characteristic of aSAH cases

| Characteristic |  | Treatment group (%) |  | Total<br>(n = 27) |
| --- | --- | --- | --- | --- |
|  |  | <12 hours<br>(n = 10) | ≥12 hours<br>(n = 17) |  |
| Sex | Female | 90.0 | 64.7 | 74.1 |
|  | Male | 10.0 | 35.3 | 25.9 |
| Age group | < 60 years | 60.0 | 41.7 | 48.1 |
|  | ≥ 60 years | 40.0 | 58.3 | 51.9 |
| WFNS at presentation | 1-3 | 60.0 | 76.5 | 70.4 |
|  | 4-5 | 40.0 | 23.5 | 29.6 |
| Interfacility transfer | Yes | 10.0 | 64.7 | 44.4 |
|  | No | 90.0 | 35.3 | 55.6 |
| Treatment type | Coiling | 70.0 | 52.9 | 59.3 |
|  | Clipping | 30.0 | 47.1 | 40.7 |
| Time to treatment | Median | 7.8 | 22.5 | 15.1 |
| (hours) | (IQR) | (6.0, 9.0) | (15.8, 26.3) | (9.0, 24.1) |

While we were unable to identify statistically significant difference in median of time to services between groups, the magnitude of the differences between certain groups was noticeable (Table 2). Men had longer time than women during pre-hospital and transfer periods. Patients requiring interfacility transfer had over 10 hours longer time from onset to definitive treatment; patients aged ≥60 years and patients treated with surgical clipping had two to three hours longer time from onset to definitive treatment.

**Table 2.** Time to treatment for different groups

| Characteristic |  | Time to treatment (hours, median (IQR)) |  |  |  |  |
| --- | --- | --- | --- | --- | --- | --- |
|  |  | Pre-hospital | ED-Diagnosis | Transfer | In-hospital | Total |
| Sex | Female | 2.0 (1.6, 4.3) | 1.6 (0.9, 2.7) | 2.3 (1.5, 4.5) | 5.6 (3.3, 13.6) | 13.5 (7.9, 22.9) |
|  | Male | 6.8 (1.8, 9.1) | 1.4 (0.7, 5.6) | 2.7 (0.7, 5.0) | 5.9 (4.6, 8.3) | 17.0 (14.0, 24.4) |
| Age group | < 60 years | 1.9 (1.6, 4.6) | 1.4 (1.1, 2.2) | 1.7 (0.8, 3.3) | 4.7 (2.4, 9.1) | 13.0 (7.6, 22.5) |
| | $\geq 60$ years | 2.2 (1.7, 7.6) | 1.8 (0.5, 4.7) | 3.2 (1.5, 4.8) | 6.0 (4.2, 11.7) | 16.4 (10.4, 25.8) |
| WFNS | 1-3 | 2.0 (1.6, 6.0) | 1.6 (1.1, 3.0) | 2.0 (0.8, 4.8) | 5.9 (3.9, 11.3) | 15.8 (9.0, 24.4) |
|  | 4-5 | 3.4 (1.7, 7.0) | 1.3 (0.5, 3.9) | 3.3 (2.0, 4.5) | 5.0 (2.2, 7.7) | 12.3 (9.3, 21.1) |
| Interfacility transfer | Yes | 2.5 (1.7, 7.9) | 1.4 (0.9, 2.7) | 2.3 (0.8, 4.8) | 13.1 (6.4, 16.0) | 19.7 (14.8, 24.7) |
|  | No | 1.9 (1.6, 5.6) | 1.6 (0.8, 4.8) | n.a. | 5.6 (3.6, 9.9) | 9.0 (6.8, 21.6) |
| Treatment type | Coiling | 2.1 (1.6, 8.0) | 2.3 (1.4, 5.4) | 1.8 (0.5, 2.2) | 5.1 (3.6, 11.0) | 13.7 (8.6, 25.0) |
|  | Clipping | 2.0 (1.6, 4.4) | 1.1 (0.6, 1.4) | 4.6 (1.7, 5.5) | 5.9 (2.8, 12.9) | 15.8 (11.8, 23.3) |

### Barriers and facilitators to more rapid treatment – mixed methods result

The results from mixed-methods data triangulation of 27 aSAH cases built on the key findings from our previous thematic analysis (Nguyen, Stirling, et al., 2021). Key barriers and facilitators across patient’s journey to treatment were presented in Figure 2. The recognition of aSAH or of a life-threatening condition (herein referred as “recognition of aSAH” theme) was the key influence on time from onset to diagnosis. The availability of resources was the key influence on whether cases received early treatment. These two themes acted as either a barrier or a facilitator for timely treatment. Good coordination between health services and professionals sped up multiple time intervals in the pathway of a significant number of patients but did not have a major impact on the time to treatment during in-hospital period. The complexity of a patient’s condition also caused delays across most time periods, but only for a few cases.

**Figure 2.**
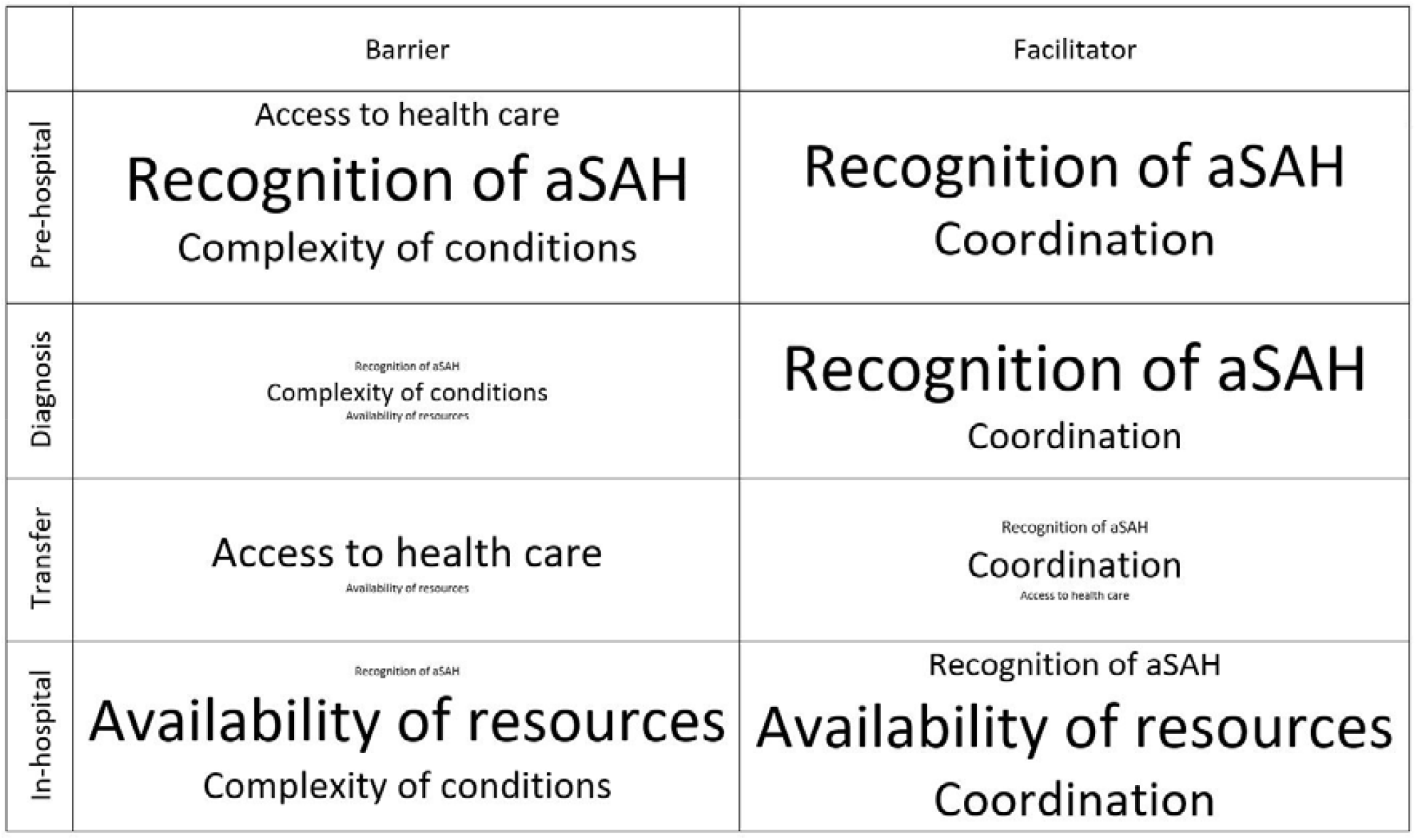
Word clouds of key barriers and facilitators across patient journey after mixed-methods triangulation

Stratifying into early and delayed treatment groups using 12 hours from onset as cut off point, we identified several distinctive factors emerging as barriers and facilitators for each treatment group for each time period (Figures 3 and 4).

**Figure 3.**
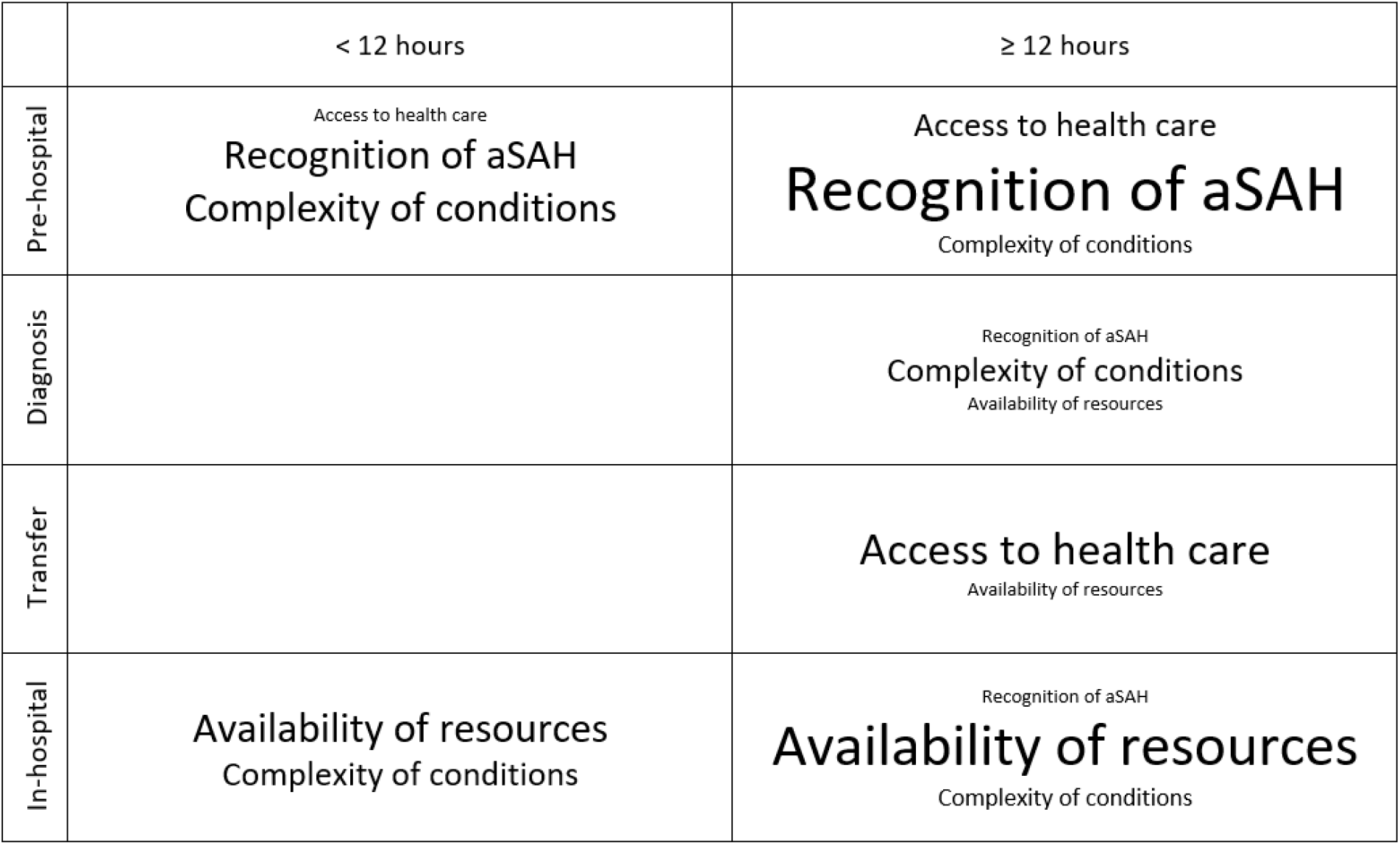
Word clouds of barriers for patients received early/delayed treatment

**Figure 4.**
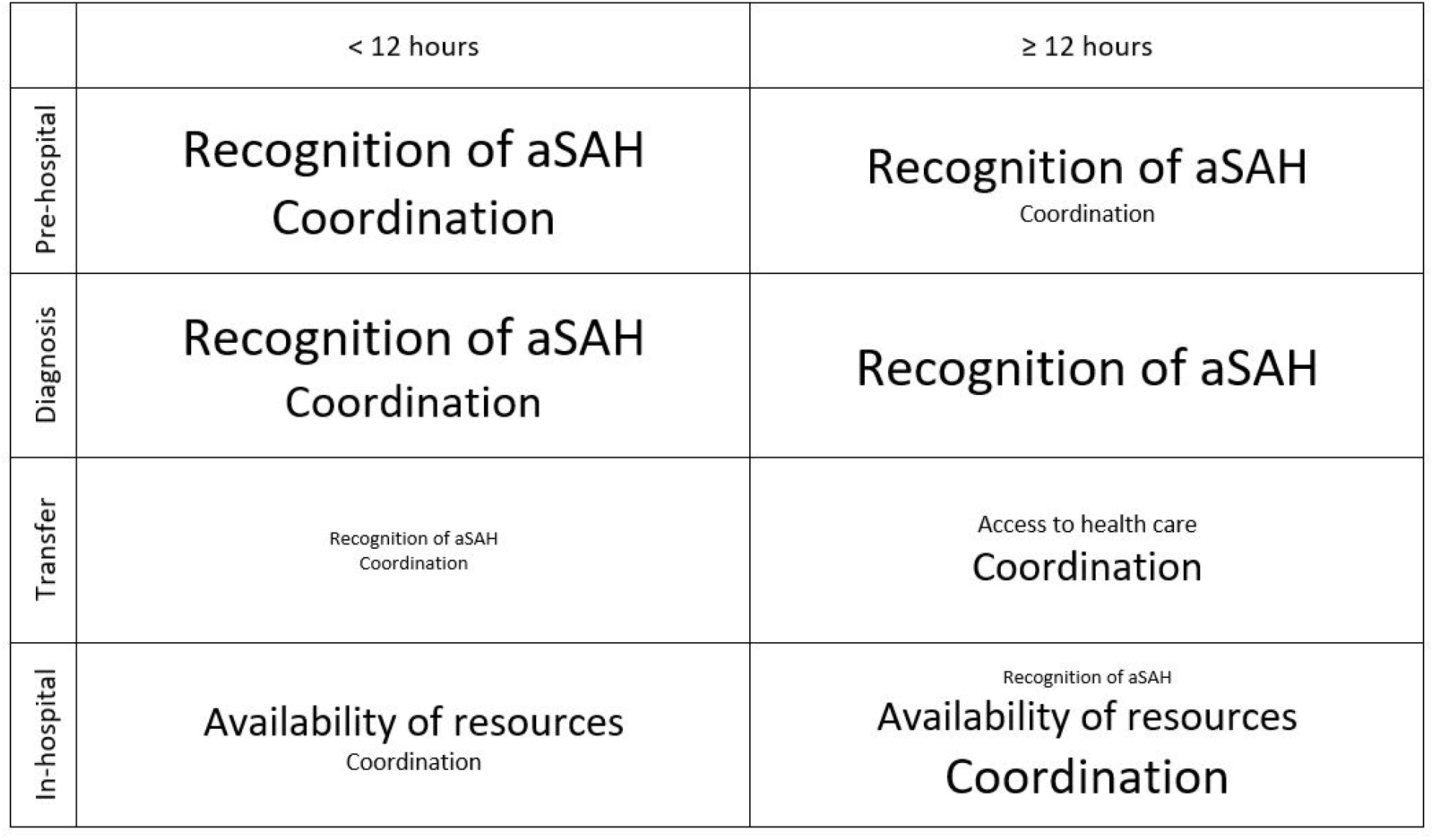
Word clouds of facilitators for patients received early/delayed treatment

#### Pre-hospital

Time duration from onset of aSAH to arrival at hospital was wide, ranging from 0.9 to 136 hours with a median of 2.1 (1.6, 7.7) hours, 1.6 (1.5, 2.0) hours for the early treatment group and 2.6 (1.7, 8.5) hours for the delayed treatment group.

“Recognition of aSAH” appeared to be the most important factor influencing timely access to health care in the pre-hospital time period. Non “recognition of aSAH” resulted in an average time of 7.8 (3.1, 10.8) hours for patients to arrive at hospital. In these instances, patients or their NOK either waited for symptoms to subside or first sought health care from their general practitioner (GP). This group tended to have a history of headache or migraine. In a small number of cases the patient was unconscious and alone.

> After that initial headache it wasn’t too bad, so he didn’t do anything about it, other than taking Panadol. And afterwards he was a little bit better, but it was still there, but he never really has a lot of headaches. […] Although he was taking a few Panadols, he just felt it’s not quite right. So, we took him to the GP in the morning. And it started from there. (Case 6 – NOK)

Early “recognition of aSAH” was the most important facilitator of timely treatment during this period. By recognising a severe, potentially life-threatening condition, patients or their NOK were able to call an ambulance or get to a hospital more rapidly with median pre-hospital time duration being 1.5 (1.4 – 1.6) hours. Even though calculation of pre-hospital time, based on data extracted from health records, was relatively short, our interview results identified seven cases where potential aSAH onset was much longer than apparent from the medical records alone. All of these patients experienced headache symptoms that were described as “sudden”, “thunderclap” or “blinding” for days or even weeks before the onset recorded in the medical records. Some of them were assessed for the headache at hospitals in emergency departments, but were not admitted during this time, and aSAH remained undiagnosed.

Paramedics also facilitated timely treatment when suspecting patients had aSAH, stroke or another neurological emergency by travelling directly to the nearest hospital with neurosurgical capacity. The strategic choice of hospital did not always directly shorten pre-hospital time because tertiary hospitals were further away than nearby regional hospitals. However, these choices had a large impact on overall time to treatment of the patients as they avoided interhospital transfer later.

> Then they put her in the ambulance and explained they were taking her to M. [tertiary hospital] rather than B. [local hospital] which would be closer to us, but they said because of her high blood pressure, could be signs of a stroke or something and this was the best place to come. (Case 4 – NOK)

Most cases used an ambulance service in our study. Generally, paramedics came within 10-15 minutes of a call which facilitated quick access to care in the pre-hospital period. However, for patients who were in hard-to-reach locations or when their condition was complicated, paramedics needed more time to retrieve or stabilise patients before transfer. This made “Access to health care” a barrier that prolonged time to hospital in four cases.

> And [the patient] had a fairly complex airway in regards that he had some vomiting, and which required two clinicians to be managing his airway in the initial stages. Initially, it was quite complex. (Case 8 – Paramedic)

“Complexity of conditions” of patients (e.g., obstructed airway, vomiting, high blood pressure, pain) was a barrier in six cases.

#### Diagnosis

Diagnosis of all aSAH occurred at the first hospital with a median time from first hospital arrival until receiving CT scan of 1.5 (0.8, 3.5) hours. The shortest diagnostic time was 0 hours while the longest time to CT was 22.3 hours. For the early treatment group, time from hospital arrival until having a CT for diagnosis was 1.3 (0.4, 0.8) hours. For the delayed group, the time period was 2.0 (1.1, 5.1) hours.

Overall, very few barriers were identified during this time period. Figure 5 presented the number of cases with each barrier/facilitator during diagnosis. Only a few cases in the delayed group experienced barriers for time to diagnosis. Patient clinical complexity caused delay to CT in three cases, for example:

**Figure 5.**
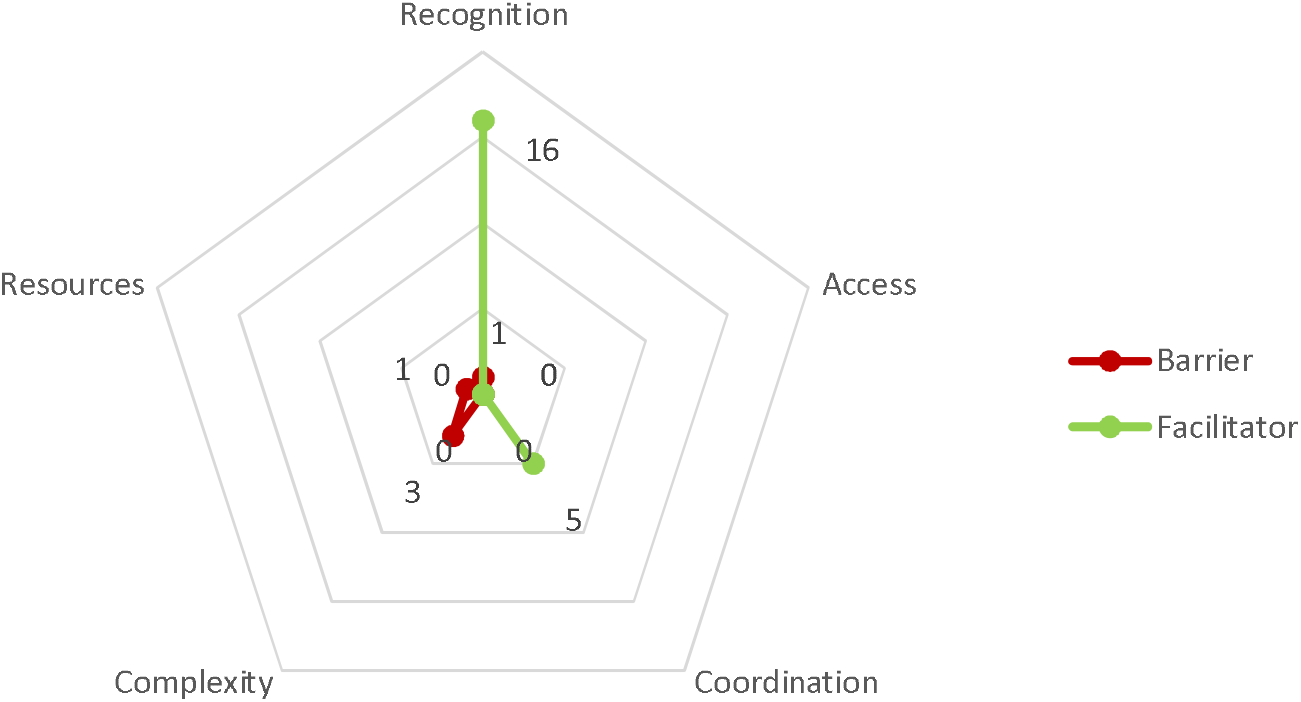
Barriers and facilitators to timely treatment during ED-diagnosis time interval

> They got an X-ray done straightaway, but they couldn’t pull her in for the CT until pretty much 24 hours after, because her respiratory rate was, I think too high, so they needed to pull that below some MET score, MET something. (Case 16 – NOK)

Only five other cases experienced delays before diagnosis with lack of recognition of aSAH in one case, lack of resources in another, and having complex conditions in three cases. The early treatment group did not encounter barriers during the diagnostic period.

Of the factors that shortened time to diagnosis “Recognition of aSAH” was dominant for both early and delayed groups. Among the early treatment group, quick recognition of a potential aSAH in the emergency department (ED) was facilitated by paramedics communication pre-hospital allowing the receiving hospital to be prepared.

> Yes, so the sequence of events with her case started with a notification from the ambulance service that they were bringing an intubated patient with a decreased conscious state, so we had some pre-notification. […]. So, we pre-notified intensive care, the stroke team and the neurosurgical registrar so that the CT scanner was available. And we actually didn’t offload her from the ambulance trolley. We just went straight to CT once we’d done a primary survey, (Case 5 – ED doctor)

We also observed that in the early treatment group, quick “recognition of aSAH” could trigger swift coordination within the hospital with faster diagnosis and early involvement of neurological specialists:

> The emergency department registrar just rang me and said - this was before they’d scanned her - he just rang and said, ‘I’ve got a lady who’s come in, in her 50s, sudden-onset headache and then rapid drop in GCS and a suspected bleed on the brain, subarachnoid haemorrhage or otherwise’. I was hanging around emergency department anyway, so I went down to the scanner and that’s where we saw what was on the scan. (Case 13 – Neurosurgeon Registrar)

For patients in the delayed treatment group, early “recognition of aSAH” at emergency departments did shorten time to diagnosis (1.1 (0.5, 1.4) hours). However, they experienced multiple barriers in the next phases that prolong overall time to treatment.

#### Transfer

Twelve patients needed interfacility transfer, with four being transferred by air ambulance and the rest transferred with ground ambulance. Median transfer time was 4.3 (2.3, 6.1) hours, with 5.0 (6.6, 7.8) hours for air transfer and 2.5 (1.7, 4.3) hours for ground transfer. Median ground distance for ground transferred patients was 21 (12.6, 171.3) km and median aerial distance for air transferred patients was 167 (160, 189.5) km. If air transferred patients had been transferred by an available ground ambulance, median driving time would have been 2.3 (2.2, 2.7) hours, as estimated using Google Maps software (Google Inc., 2021) for door-to-door distance between exact street addresses of emergency departments.

There was only one transferred case among the early treatment group. There were no barriers identified during transfer for this patient. Among 11 cases requiring transfer in the delayed treatment group, the two most important factors influencing the duration of transfer phase were “access to health care” and “coordination”.

“Access to health care” was a noticeable barrier to quicker transfer time for six cases. Longer transfer time was due to external factors including long distances between health facilities and/or bad weather conditions preventing air transfer even if the cases were assigned for urgent transfer.

> […] and then on route here they called to say they couldn’t land because of the fog. So, they had diverted to a football field near there, and then they had come by road to [the hospital]. They arrived, from memory still past midday, so there was quite a delay in getting the patient here. (Case 7 – Neurosurgeon Registrar)

We identified “lack of resources” as a transfer time barrier for one case, for which no ambulance was immediately available at the time of transfer. However, once the ambulance became available, the patient with aSAH was prioritised for transfer.

> […] for the aneurysm patient in D. and the acute subdural in D., there was only one ambulance between those three patients. So, they had to prioritise which one and [the INR] said, ‘Let’s get the aneurysm across first, that’s the one we can potentially help the most, probably’. (Case 12 – Neurosurgeon Registrar)

Smooth “coordination” between referring and receiving hospitals was a facilitator in the interhospital transfer phase. However, as in the pre-hospital phase, smooth coordination did not necessarily reduce transfer time. Instead, good coordination and early communication between health care facilities and transferring of diagnostic results sped up the treatment process at the receiving hospitals.

> Yeah, so the neurosurgical registrar … He was arranging for the bed to be requested on the ward, and once that had happened, then I got a text message back saying that the bed had been requested, and the patient was to go straight to the ward, and to request for an ambulance transfer. (Case 1 – ED doctor)

#### In-hospital

Excluding transfer time for interfacility-transferred patients, in-hospital time was 5.7 (3.6, 11.3) hours. The early treatment group had significantly shorter time (3.6 (2.5, 4.4) hours) in treating hospital than the delayed treatment group (9.1 (5.6, 15.1) hours). All themes other than ‘access to health care’ had influence on the in-hospital time period but “availability of resources” and “coordination” were the most prominent (Figure 6).

**Figure 6.**
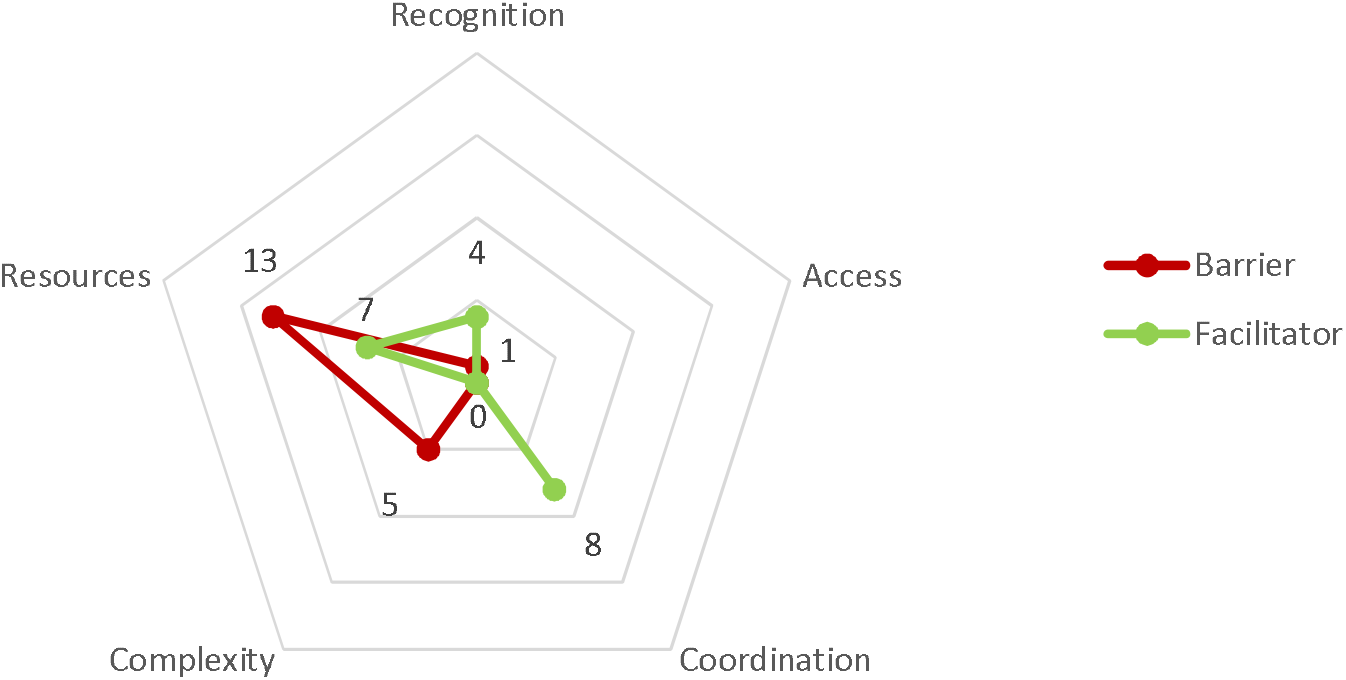
Barriers and facilitators to timely treatment during in-hospital time interval

A lack of resources was the most important barrier to quicker time to treatment. For the delayed group, a majority arrived at the hospital outside of routine hours (6am to 10pm), for which they often had to wait until the next morning for treatment. Both treating hospitals in our study consider endovascular coiling and surgical clipping as procedures that are preferentially done during regular daytime hours to avoid potential undesirable outcomes for patients as well as negative effect on the medical team.

> If the patient were in extremes [critical], then of course there’s flexibility of that, but typically because of the complexity of the procedure, it’s not something that you want to do really, really late. So, if by our reckoning we’re not going to be able to get started by 10 [pm], then it’s usually safer to just do it first thing in the morning. (Case 16 – Interventional Neuroradiologist)

The early treatment group were also affected by the unavailability of resources but mainly due to limited capacity of operating theatres and/or availability of neurosurgeons or interventional neuroradiologists. These cases could not receive treatment immediately but unlike the delayed group, they did not have to wait for an extended duration overnight.

> At that stage there were two theatres running, so an anaesthetist wasn’t going to be available for another hour-and-a-half or so, so a plan was made to bring a team in to do the case, endovascular in about an hour-and-a-half. (Case 11 – Interventional Neuroradiologist)

Smooth coordination between facilities, as well as within a treating facility, played an important role in speeding up the treatment process for patients. When a patient was in transit prior preparation at the tertiary hospital was crucial to ensure patient could rapidly go into treatment if resources allowed. If it was impossible for them to receive treatment due to resource constraints, their treatment was arranged for first thing the following morning.

> So, I think pretty much the night before, the neurosurgery reg on call, had been liaising with anaesthetics and they said they were pretty happy to do it first case, unless some other emergency comes up, which fortunately didn’t. So, we were able to pretty much proceed as planned. (Case 3 – Interventional Neuroradiologist)

‘Complexity of conditions’ was a barrier to rapid definitive treatment in five cases (three in the early treatment group and two in the delayed treatment group). In all cases, the patients’ conditions required additional stabilising procedures before they could receive endovascular coiling or surgical clipping.

## Discussion

The ‘Diamond Approach’ provided clarity about the key factors influencing timely treatment of aSAH. Early recognition of aSAH or severe condition, good coordination during pre-hospital and diagnosis phases, and availability of resources for treatment during the in-hospital period were critical factors in receiving treatment within 12 hours of aSAH onset. In contrast, lack of recognition of aSAH at onset and lack of resources for immediate in-hospital treatment were major factors in delaying treatment beyond 12 hours. Initial thematic analysis of qualitative data identified five key themes. Incorporating quantitative data on time to key events in the treatment pathway for each case and subsequent categorisation of participants as early or late treatment enabling further refinement of qualitative analysis to three key factors that had major impacts on time to treatment. Using a word cloud analysis and data presentation technique provided further clarity for analysis and display.

Our study consisted of a sample of 27 patient cases with aSAH purposefully sampled to maximise the diversity of patients’ socio-economic backgrounds across two tertiary referral networks. Patient characteristics in our sample were similar to those from other aSAH studies: the proportion of female, of lower WFNS grade (1-3), and type of treatment (coiling/clipping) were all similar (Baltsavias et al., 2000; Dupont et al., 2009; Nichols et al., 2016; Rehman et al., 2020), suggesting our sample adequately reflects aSAH patients.

“Recognition of aSAH” appears to be the most important facilitator and barrier to timely treatment of aSAH, a finding that is supported by the work of Kassell et al (1985) who identified that unspecific symptoms of aSAH frequently led to the misrecognition of an aSAH event (Kassell et al., 1985). It also appears that aSAH may be unrecognised for an extended time period (e.g., days to weeks) if symptoms are believed to be benign headache or migraine. Evidence for this came from interviews and conflicted with the information in the medical record. It is therefore possible that administrative datasets normally used for study of aSAH may not capture this time period well. Key sign to distinguish the clinical characteristics of headache in nontraumatic aSAH with other types of headache remain limited. Headache is one of the most common reasons for presentation to hospital ED for people <60 years, about one-fifth of them experienced instant peak and one-third described it as “worst ever” headache in their life and yet the incidence of aSAH is very low, around 3% (Chu et al., 2017). However, a rapidly intensifying headache (e.g., seconds to a minute) that becomes “the worst headache of someone’s life” is the most frequent description for aSAH (Linn et al., 1998; Mac Grory et al., 2018; Perry et al., 2010). Additional symptoms in these studies such as a stiff neck, vomiting, a witnessed loss of consciousness or raised blood pressure are also symptoms more likely to be experienced by aSAH patients presenting with headache compared to headache in patients without aSAH. Hence, these signs could be communicated to the community to raise awareness of potentially severe headaches that need emergency care. The intensity of headache and the time for the headache to reach its peak should be key features of the patients’ history to identify potential aSAH by health professionals. More research to understand differentiating features of milder headaches associated with aSAH could improve patient outcomes by improving early aSAH recognition. A detailed medical history from any potential aSAH patients to identify the likely onset of event could facilitate more appropriate triage and better diagnosis for patients.

Good coordination was identified as an overarching facilitator for more timely treatment across multiple time periods. Good coordination started when patients first contacted the health system and continued until patients received definitive treatment. Communication between the interprofessional team is most crucial for effective care coordination (Dingley et al., 2008). Receiving pre-notification of arriving patients with urgent conditions such as stroke or trauma has been shown by others to shorten time spent in emergency departments (Handolin & Jääskeläinen, 2008; Patel Mehul et al., 2011). Intrahospital and interhospital transfer also require effective communication and interaction between departments/facilities to improve patient transfer workflow (Abraham & Reddy, 2010). Our results support the idea that patient transfer procedure guidelines and effective information technologies can facilitate the transfer process (Abraham & Reddy, 2010; Sanner & Øvrelid, 2020). Communication between facilities during the pre-hospital and transfer periods did not directly reduce these time periods but resulted in more timely treatment at the treating hospitals. Clear and detailed guidelines on workflow for patient with aSAH along with knowledgeable health professionals facilitated timely management of aSAH cases.

There were few barriers or facilitators for timely treatment during the diagnostic and transfer time periods for patients in our study. Similar to other studies, having interfacility transfer contributed to the total time from onset to definitive treatment (Nichols et al., 2020; Weyhenmeyer et al., 2018). However, the effect of air transfer on time to treatment remains ambiguous. Others have found that for a similar distance, air transfer took more (Paoli et al., 2020; Regenhardt et al., 2018) or less (Svenson et al., 2006) time compared to ground transfer. In our study, cases with air transfer had longer transfer time than ground transferred cases, but they were much further away from the treating hospital. Air transfer is highly dependent on weather conditions as identified in our study. Nocturnal air transfer has also been proved to increase delay in transfer for stoke patients (Regenhardt et al., 2018). Hence, mode of transport should be critically considered based on distance, weather conditions and time of the day.

Availability of resources was a crucial determinant of timely treatment. Even when a patient was diagnosed and transferred quickly to a neurosurgical unit, when this occurred during the night, or when health specialists or theatres were unavailable, patients could not receive immediate treatment. While there is a large body of evidence showing an increased risk of mortality from night-shift surgery the certainty of such evidence is low (Cortegiani et al., 2020). In contrast, risk of early mortality due to aSAH comes mainly from rebleeding of the ruptured aneurysm (Larsen & Astrup, 2013), with the risk increasing with length of duration since onset (van Donkelaar Carlina et al., 2015). Therefore, better evidence about the balance of risk from night-shift surgery and rebleeding on outcomes after aSAH is needed.

### Contribution to the field of mixed methods

We used an adapted convergent mixed methods multiple case study design in which quantitative and qualitative data were collected, analysed and interpretated in six steps (Creswell & Creswell, 2018). Case studies are used to obtain a comprehensive understanding of a phenomenon or an event in its natural real-life setting (Crowe et al., 2011) and are useful for health services researchers wanting to explore an issue from different perspectives (Yin, 2018). With the aim of understanding aSAH events from the perspective of different stakeholders while identifying potential causes for delays, a multiple case study approach was a suitable method to answer the research question. Multiple cases allow observation of replicating patterns, thereby increasing the robustness of the findings. Since case studies rely on analytical rather than statistical generalisations, relying on replication logic provided external validation to the findings. Each case served to confirm or disconfirm the conclusions drawn from the others (Yin, 2018).

Triangulation has been built in every stage of our study. We employed methods triangulation where we combined both qualitative and quantitative methods to collect complementary data on treatment of aSAH (Carter et al., 2014). Investigator triangulation was used by having at least two investigators to collect, code, analyse and interpret data to bring both confirmation of findings and to add breadth to the phenomenon of interest (Denzin & Lincoln, 2005). The case study approach allowed us to triangulate data sources by collecting data from different stakeholders on the same phenomenon to gain multiple perspectives and validation of data (Carter et al., 2014). Our Diamond method allowed us to synthesise and triangulate the data both within and across cases. By coding within cases blinded from treatment status, we could minimise bias in identify potential barriers and facilitators of timely treatment in Step 1 of The Diamond Approach. Next, we were able to triangulate the facilitators and barriers identified in each period with the actual time duration. Quantitative measurements added precision to interview information, strengthening our findings and allowing us to clarify and validate the major facilitators and barriers to timely treatment for each phase during the patient’s pathway. At the same time, the richness of information from the interviews provided insights into what happened during particular time periods, enriching understanding of the dynamic interaction of factors in the diverse and complex settings of each aSAH case.

Our method increases the validity and trustworthiness of qualitative data through achieving credibility, transferability, dependability, and confirmability (Nowell et al., 2017). Data were collected and triangulated from multiple sources ensuring the internal validity of the research, which therefore increases credibility. Transferability means that study findings could be applicable to other similar situations and contexts. To achieve transferability, one common solution is to do purposive sampling to maximise the data regarding a certain context (Shenton, 2004). Our study has employed such an approach by purposively selecting patients with diverse socio-economic backgrounds to maximise data richness and diversity. Dependability means that with the same design, tools and context, the same results could be produced by good, logical and traceable documentation. We used “Codebook” thematic analysis approach which allowed clear documentation of the coding process (Braun & Clarke, 2020). We created a unified database consisting of all qualitative and quantitative data in NVivo, allowing all data analysis traceable and documented. Confirmability is about how conclusions and interpretations must be derived from the data collected once credibility, transferability and dependency are achieved (Nowell et al., 2017).

### Limitations

Our study has several limitations. First, while we interviewed 90 key informants, we could not recruit all key health professionals involved in each case. Hence, in several cases where patients experienced delays in a certain time interval, we could not interview health professionals in-charge to comprehensively understand causes of delays. However, by interviewing at least two key informants for each case, we minimised such information gaps. Second, with the convergent approach in which qualitative and quantitative data are collected in parallel, we were unable to verify patients’ time to treatment prior to interview with health professional to facilitate their reflections on causes of delays.

## Conclusion

In conclusion, our approach to data analysis and synthesis in a mixed methods multiple case study (The Diamond Approach) advanced the convergent mixed-methods design and provides guidance for data synthesis for future clinically focused mixed methods studies. Using The Diamond Approach, we identified the most significant factors affecting treatment within 12 hours from onset of aSAH, including early recognition of aSAH during pre-hospital and diagnosis stages, proper coordination across all stages, and availability of resources to conduct definitive treatment during in-hospital time. Improving recognition of aSAH or recognition of a severe event for the general population could facilitate earlier seeking of emergency medical care. More thorough patient history-taking and headache examination at ED could also help to identify potential aSAH cases. Although aSAH symptoms are often nonindicative, the unbearable intensity and short time to peak could be an indicator for suspecting aSAH. Availability of resources for treatment of aSAH can be considered through healthcare system design to help achieve more favourable outcomes for patients.

## Data Availability

Due to the nature of this research, participants of this study did not agree for their data to be shared publicly, so supporting data is not available.

