## Supplement for "Advancing convergent mixed methods design using the Diamond Approach for clinical multiple case studies: an example using data on time to treatment in aneurysmal subarachnoid haemorrhage"

Supplement 1 – Interview tree

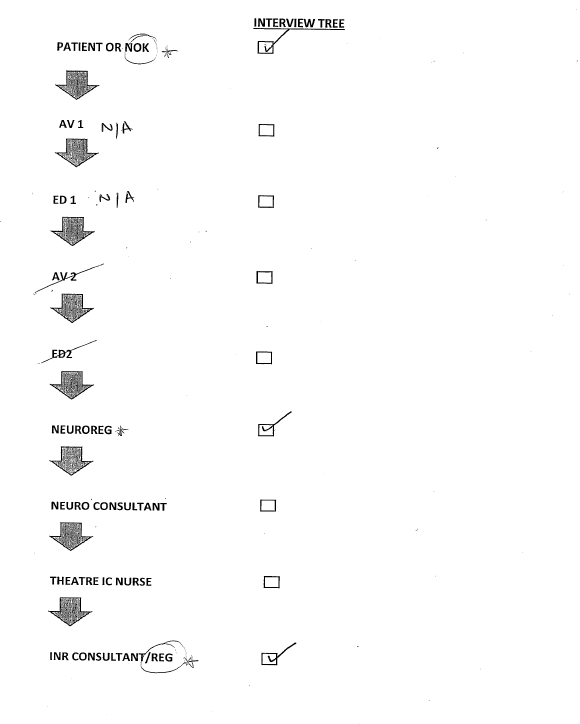

Note: NOK: Next-of-kin; AV: Ambulance staff; ED: Emergency doctor; REG: registrar; INR: Interventional Neuroradiology; IC: internsive care

Supplement 2 – Summary table of all cases

| Case | Pre-hospital | Diagnosis | Transfer | In-hospital | Total |
| --- | --- | --- | --- | --- | --- |
| (1)  F, 80+  Transfer  Coiling | 2.6  **Recognition (↓)** | 2.0  Recognition (↓)  **Complexity (↑)** | 1.8 | 8.7  **Lack of resources (↑)** | 15.1  Potential onset few days earlier (from interview) |
| (2)  F, <50  Non-transfer  Clipping | 1.6  **Recognition (↑)** | 2.2  Recognition (↓)  **Complexity (↑)** |  | 9.1  **Complexity (↑)**  Good coordination (↓) | 13.0 |
| (3)  F, 60-69  Transfer  Coiling | 8.5  **Recognition (↑)**  **Access (↑)** | 0.5  **Recognition (↓)** | 4.2  **Access (↑)**  Good coordination (↓) | 13.1  **Lack of resources (↑)**  Good coordination (↓) | 26.3  No clear onset time |
| (4)  F, 50-59  Non-transfer  Coiling | 0.9  **Recognition (↓)**  Good coordination (↓) | 1.9 |  | 2.4  **Availability of resources (↓)** | 5.2 |
| (5)  F, <50  Non-transfer  Clipping | 1.9  **Recognition (↓)**  **Good coordination (↓)** | 0.1  **Recognition (↓)**  **Good coordination (↓)** |  | 1.0  **Availability of resources (↓)** | 3 |
| (6)  M, 70-79  Non-transfer  Coiling | 136.0  **Recognition (↑)** | 6.8  **Recognition (↑)** |  | 12.1  **Complexity (↑)**  Availability of resources (↓) | 155.0  No clear onset time |
| (7)  F, <50  Transfer  Clipping | 7.6  **Recognition (↑)** | 1.0 | 5.3  **Access (↑)** | 0.0  **Good coordination (↓)** | 14.0  No clear onset time |
| (8)  M, 70-79  Non-transfer  Coiling | 6.8  **Recognition (↑/↓)**  **Good coordination (↓)**  **Complexity (↑)** | 0.0  **Recognition (↓)**  **Good coordination (↓)** |  | 2.9  **Availability of resources (↓)** | 9.8  No clear onset time |
| (9)  F, 60-69  Non-transfer  Coiling | 1.4  **Recognition (↓)** | 3.5 |  | 4.1  **Availability of resources (↓)** | 9.0 |
| (10)  F, 50-59  Transfer  Clipping | 1.5  **Recognition (↓)** | 1.4  **Recognition (↓)** | 4.5 | 16.6  **Lack of resources (↑)**  Good coordination (↓) | 24.0  Potential onset few days earlier (from interview) |
| (11)  F, 60-69  Non-transfer  Coiling | 2.1  **Recognition (↑)**  **Complexity (↑)** | 0.4  **Recognition (↓)** |  | 3.6  **Lack of resources (↑)**  **Complexity (↑)** | 6.0  Potential onset few days earlier (from interview) |
| (12)  F, <50  Transfer  Clipping | 4.6  **Recognition (↑)** | 3.5 | 2.5  **Lack of resources (↑)**  **Recognition (↓)**  **Good coordination (↓)** | 0.1  **Recognition (↓)**  **Good coordination (↓)** | 10.7  No clear onset time |
| (13)  F, 50-59  Non-transfer  Coiling | 1.6  **Recognition (↑)**  **Complexity (↑)**  **Access (↑)**  **Good coordination (↓)** | 1.5  **Recognition (↓)**  **Good coordination (↓)** |  | 4.5  **Lack of resources (↑)**  **Complexity (↑)** | 7.6  Potential onset few days earlier (from interview) |
| (14)  F, 80+  Transfer  Coiling | 1.7  **Recognition (↓)**  **Complexity (↑)** | 5.1 | 0.5 | 15.1  **Lack of resources (↑)** | 22.5  Potential onset few days earlier (from interview) |
| (15)  M, 70-79  Transfer  Coiling | 1.2  **Recognition (↑)** | 5.1 | 1.5  **Good coordination (↓)** | 4.5  **Recognition (↓)**  **Availability of resources (↓)**  **Good coordination (↓)** | 12.3 |
| (16)  F, 50-59  Non-transfer  Coiling | 11.2  **Recognition (↑/↓)** | 22.3  Recognition (↓)  **Complexity (↑)** |  | 17.5  **Lack of resources (↑)** | 51.0  No clear onset time |
| (17)  F, <50  Non-transfer  Coiling | 1.4  **Recognition (↓)**  **Good coordination (↓)** | 1.1  **Recognition (↓)**  **Good coordination (↓)** |  | 3.6  **Lack of resources (↑)**  **Complexity (↑)** | 6.1  Potential onset few days earlier (from interview) |
| (18)  F, <50  Non-transfer  Clipping | 1.6  **Recognition (↓)**  **Complexity (↑)**  **Good coordination (↓)** | 0.5  **Recognition (↓)**  **Good coordination (↓)** |  | 5.9  **Recognition (↓)**  **Good coordination (↓)** | 8.1 |
| (19)  M, 80+  Non-transfer  Coiling | 7.8  **Recognition (↑)** | 6.2 |  | 10.6  **Lack of resources (↑)** | 24.6 |
| (20)  F, 50-59  Non-transfer  Coiling | 2.2  **Recognition (↓)**  **Complexity (↑)**  Good coordination (↓) | 12.9  **Lack of resources (↑)** |  | 5.6  **Recognition (↑)** | 20.6 |
| (21)  F, <50  Non-transfer  Clipping | 4.3  **Recognition (↑)** | 1.4  **Recognition (↓)** |  | 16.9  **Lack of resources (↑)** | 22.5 |
| (22)  F, 60-69  Transfer  Clipping | 2.3  **Recognition (↓)**  **Access (↑)** | 0.6  **Recognition (↓)** | 5.6  **Access (↑)** | 32.1  **Lack of resources (↑)** | 40.6 |
| (23)  M, 60-69  Transfer  Clipping | 1.7  **Good coordination (↓)** | 0.3  **Recognition (↓)** | 7.8  **Access** **(↑)**  Recognition (↓)  Good coordination (↓) | 6.1  **Lack of resources (↑)** | 15.8 |
| (24)  F, 60-69  Transfer  Coiling | 49.4  **Recognition (↑)** | 2.5 | 3.8  **Access (↓)**  **Good coordination (↓)** | 2.6  **Availability of resources (↓)**  **Good coordination (↓)** | 58.4  No clear onset time |
| (25)  F, 70-79,  Non-transfer  Coiling | 1.6  **Recognition (↓)** | 1.6  **Recognition (↓)** |  | 5.7  **Lack of resources (↑)** | 9.0  Potential onset few days earlier (from interview) |
| (26)  M, 60-69  Transfer  Clipping | 2.0  **Recognition (↑)** | 1.1  **Recognition (↓)** | 8.0  **Access (↑)** | 5.9 | 17.0  No clear onset time |
| (27)  M, 50-59  Transfer  Clipping | 10.4  **Recognition (↑)**  **Access (↑)** | 1.4 | 7.8  **Access (↑)** | 4.7  **Recognition (↓)** | 24.2  No clear onset time |

- **Bold: Primary factor**
- Non-bold: Secondary factor
- ↑: Barrier (increase time to treatment)
- ↓: Facilitator (shorten time to treatment)
